# New-onset prostate cancer in type 2 diabetes mellitus exposed to the SGLT2I, DPP4I and GLP1a: A population-based cohort study

**DOI:** 10.1101/2023.11.25.23298886

**Authors:** Oscar Hou In Chou, Lei Lu, Cheuk To Chung, Jeffrey Shi Kai Chan, Raymond Ngai Chiu Chan, Athena Lee Yan Hiu, Edward Christopher Dee, Kenrick Ng, Hugo Hok Him Pui, Sharen Lee, Bernard Man Yung Cheung, Gary Tse, Jiandong Zhou

**Author notes:** Co-first author (Oscar Hou In Chou, Lei Lu). Correspondence to: Jiandong Zhou, Ph.D., Division of Experimental Medicine, Nuffield Department of Medicine, University of Oxford, Oxford, United Kingdom.

## Abstract

**Background:** Sodium-glucose cotransporter 2 inhibitors (SGLT2I) have been suggested to reduce new-onset cancer amongst type-2 diabetes mellitus (T2DM) patients.

**Objective:** This real-world study aims to compare the risks of prostate cancer between SGLT2I and dipeptidyl peptidase-4 inhibitors (DPP4I) amongst T2DM patients.

**Design, setting and participants:** This was a retrospective population-based cohort study of prospectively recorded data on type-2 diabetes mellitus (T2DM) male patients prescribed either SGLT2I or DPP4I between January 1^st^ 2015 and December 31^st^ 2020 from Hong Kong.

**Methods:** The primary outcome was new-onset prostate cancer. The secondary outcomes included cancer-related mortality and all-cause mortality. Propensity score matching (1:1 ratio) using the nearest neighbour search was performed and multivariable Cox regression was applied to compare the risk. A three-arm sensitivity analysis including the glucagon-like peptide-1 receptor agonist (GLP1a) cohort was conducted.

**Results:** This study included 42129 male T2DM patients (median age: 61.0 years old [SD: 12.2]; SGLT2I: n=17120; DPP4I: n=25009). After matching, the number of prostate cancers was significantly lower in SGLT2I users (n = 60) than in DPP4I (n = 102). SGLT2I use was associated with lower prostate cancer risks (HR: 0.45; 95% CI: 0.30-0.70) after adjustments than DPP4I. The results remained consistent in the sensitivity analysis. SGLT2I reduced the risks of prostate cancer prominently amongst patients who were older (age >65), patients with 2^nd^ and 3^rd^ quartile of HbA1c, concurrent metformin uses, and concurrent sulphonylurea uses. SGLT2I was associated with higher risks of prostate cancer amongst sulphonylurea non-users.

**Conclusion:** The real-world study demonstrated SGLT2I was associated with lower risks of new-onset prostate cancer after matching and adjustments compared to DPP4I. This result warrants further prospective studies.

**Graphical abstract:** 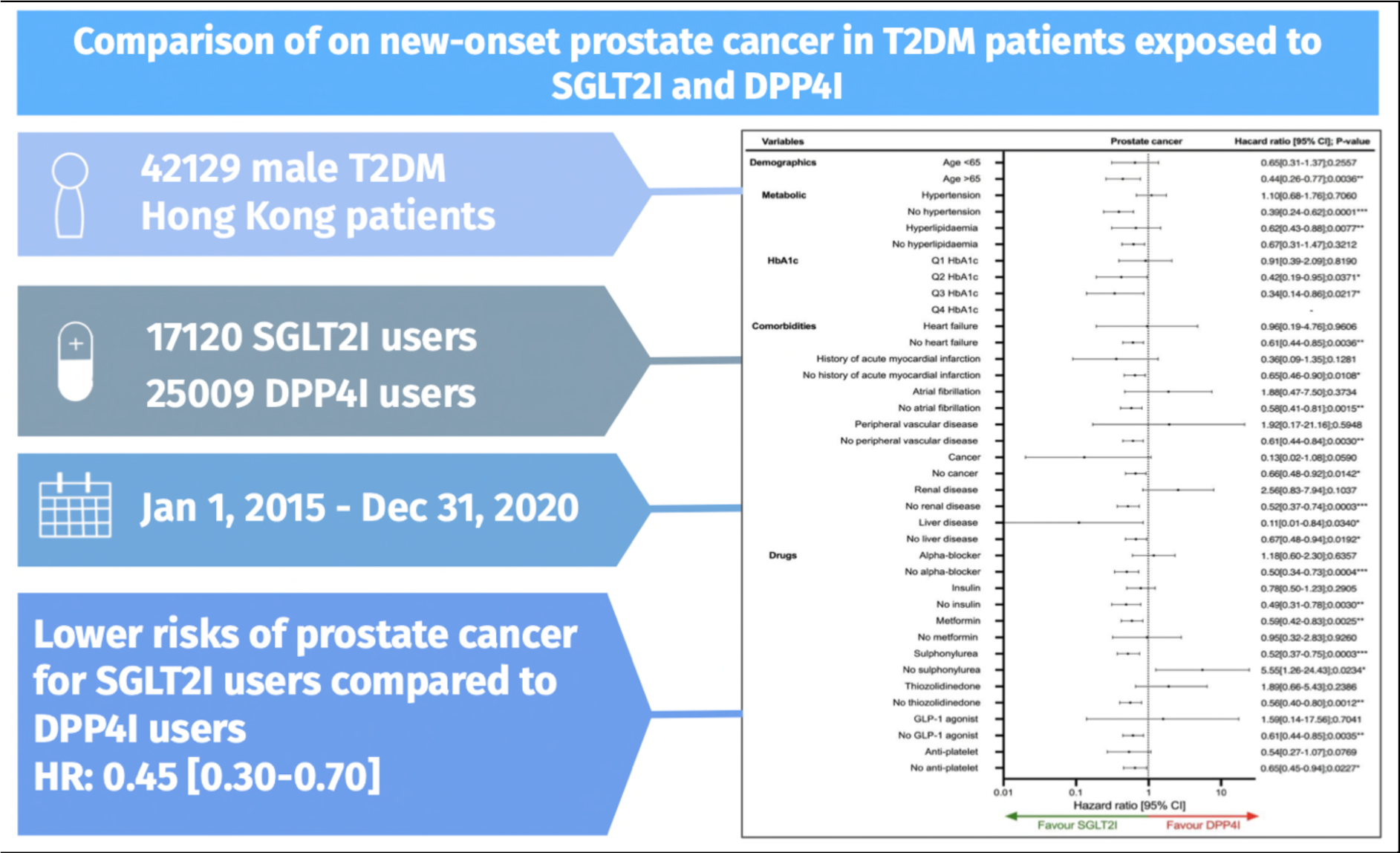

## Introduction

Type 2 diabetes mellitus (T2DM) and prostate cancer are two of the most common chronic diseases that are affected by the ageing male population [1, 2]. Despite relatively lower rates of prostate cancer in Asia compared to Northern America, there have been increasing incidence rates in several Asian countries, which is potentially related to the rising prevalence of obesity, metabolic aberrations and T2DM [3, 4]. The current literature suggests an association between T2DM and cancer proliferation, however its influence on the development of prostate cancer remains controversial [5]. This notwithstanding, there is also evidence suggesting that SGLT2I demonstrate a protective effect against new-onset overall cancer compared to DPP4I [6]. This has sparked interest in inquiring whether better diabetic management can reduce the risk of prostate cancer via existing anti-diabetic medications. Subsequently, this may allow clinicians to consider alternative treatment options for those at risk of prostate cancer.

Previous results regarding the association of prostate cancer and metformin use remains controversial [7–10]. This further propagated research on novel anti-diabetic agents such as sodium-glucose cotransporter 2 inhibitors (SGLT2I) and dipeptidyl peptidase 4 inhibitors (DPP4I) on prostate cancer; as these drugs are the novel second line anti-diabetic medication commonly prescribed in Hong Kong and other developed countries. Previous studies has shown that the inhibition of SGLT2-mediated glucose uptake could reduce the proliferation of prostate cells molecularly [11]. Meanwhile, the clinical data on SGLT2I remained scarce. For direct comparison between the SGLT2I and DPP4I usage in the clinical setting, a study found no significant difference in risk of prostate cancer [12]. As of now, there is limited clinical evidence surrounding the association of SGLT2I and DPP4I on prostate cancer. This present study aims to investigate the role of SGLT2I and DPP4I with new-onset prostate cancer in a cohort of T2DM patients from Hong Kong.

## Methods

### Study population

This study was approved by the Institutional Review Board of the University of Hong Kong/Hospital Authority Hong Kong West Cluster (HKU/HA HKWC IRB) (UW-20-250) and complied with the Declaration of Helsinki. This was a retrospective population-based study of prospectively collected electronic health records using the Clinical Data Analysis and Reporting System (CDARS) by the Hospital Authority (HA) of Hong Kong. The records cover both public hospitals and private clinics in Hong Kong. It was verified that over 90% of the identified T2DM patients were under the HA [13, 14]. This system has been used extensively by our teams and other research teams in Hong Kong. The system contains data on disease diagnosis, laboratory results, past comorbidities, clinical characteristics, and medication prescriptions. The system has been used by our Hong Kong research team to perform comparative studies [9, 15, 16]. T2DM patients who were administered with SGLT2I or DPP4I in centres under the Hong Kong HA, between January 1st, 2015, to December 31st, 2020, were included. The glucagon-like peptide-1 receptor agonist (GLP1a) cohort was included for sensitivity analysis to demonstrate the relative effects amongst the second line oral anti-diabetic agents.

### Predictors and variables

Patients’ demographics include gender and age of initial drug use (baseline), clinical and biochemical data were extracted for the present study. Prior comorbidities were extracted in accordance with the *International Classification of Diseases Ninth Edition* (ICD-9) codes (**Supplementary Table 1**). The diabetes duration was calculated by examining the earliest date amongst the first date of (1) diagnosis using of ICD-9; (2) Hba1c >=6.5%; (3) Fasting glucose >= 7.0 mmol/l or Random glucose 11.1 mmol/l 4) Using anti-diabetic medications. The number of hospitalisations in the year prior the index days were extracted. The Charlson’s standard comorbidity index was calculated [17]. The prostate medication (alpha-blockers), cardiovascular medications, anti-diabetic agents, and were extracted from the database. The duration and frequency of SGLT2I and DPP4I usage were calculated. The baseline laboratory examinations, including the glucose profiles and renal function tests were extracted. The microbiology testing results were also extracted to define the urinary tract infection **(Supplementary Table 1)**. The estimated glomerular filtration rate (eGFR) was calculated using the abbreviated modification of diet in renal disease (MDRD) formula [18]. The following patient groups were excluded: those who (1) with prior prostate cancer (n=135); (2) died within 30 days after initial drug exposure (n=167); (3) without complete demographics (n=11); (4) under 18 years old (n=29); (5) new onset prostate cancer development less than 1 year after drug exposure (n=20) (**Figure 1**). To account for the incomplete laboratory data, the multiple imputation by chained equations was performed according to the previous published study [19].

**Figure 1.**
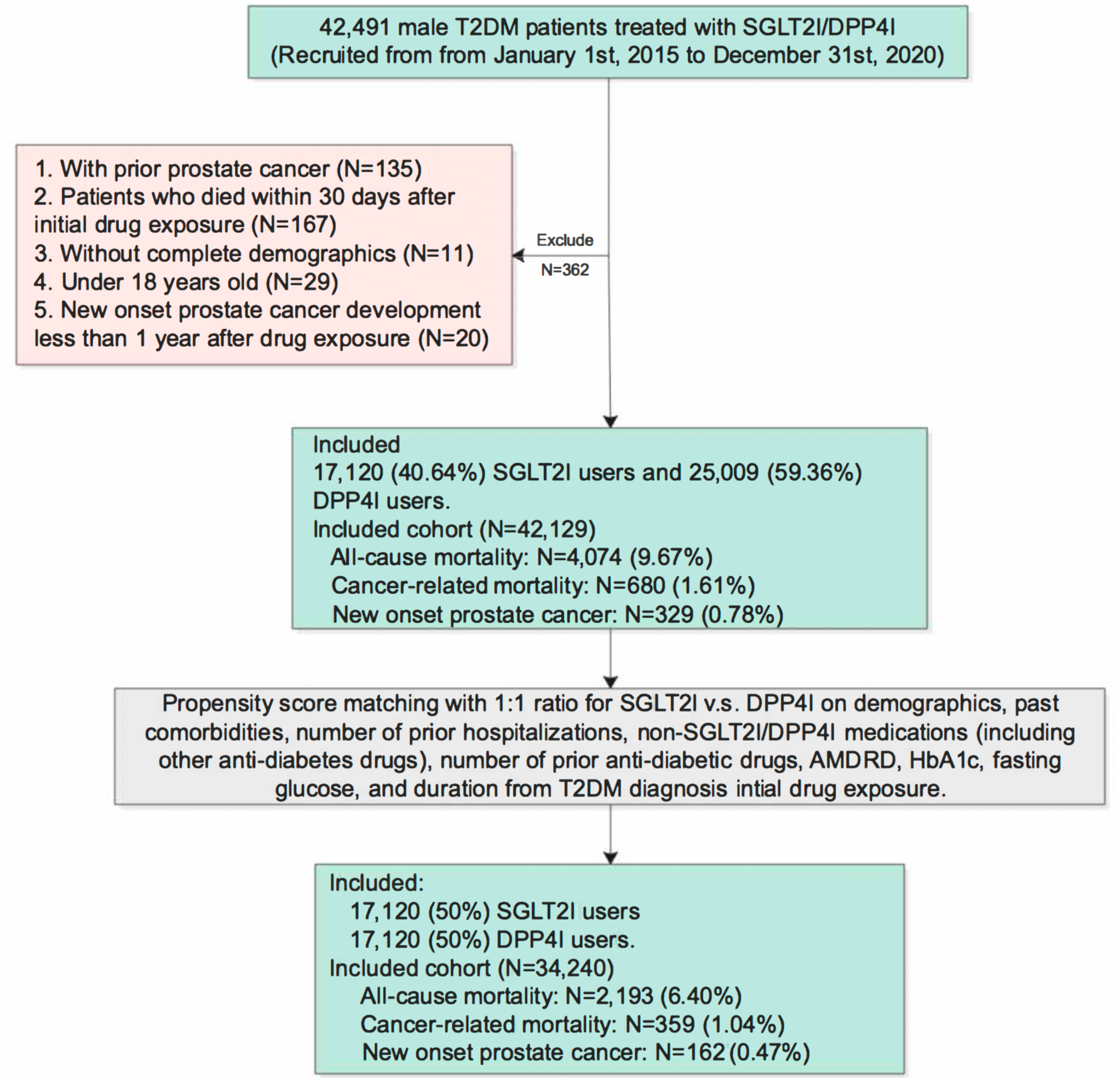
Procedures of data processing for the study cohort. SGLT2I: Sodium-glucose cotransporter-2 inhibitors; DPP4I: Dipeptidyl peptidase-4 inhibitors. MDRD: modification of diet in renal disease.

### Study outcomes

The primary outcome of this study was new-onset prostate cancer (ICD9: 185) after the index date of the drug use. The secondary outcomes were cancer-related mortality and all-cause mortality. Mortality data were obtained from the Hong Kong Death Registry, a population-based official government registry with the registered death records of all Hong Kong citizens linked to CDARS. Mortality was recorded using the *International Classification of Diseases Tenth Edition* (ICD-10) coding. The as-treat approach was adopted which patients were censored at treatment discontinuation or switching of the comparison medications. The endpoint date of interest for eligible patients was the event presentation date. The endpoint for those without primary outcome was the mortality date or the end of the study period (31^st^ December 2020).

### Statistical analysis

Descriptive statistics are used to summarize baseline clinical and biochemical characteristics of patients with SGLT2I and DPP4I use. For baseline clinical characteristics, continuous variables were presented as mean (95% confidence interval/standard deviation [SD]) and the categorical variables were presented as total numbers (percentage). Propensity score matching generated by logistic regression with 1:1 ratio for SGLT2I use versus DPP4I use based on demographics, number of prior hospitalization, non-SGLT2I/DPP4I medications, number of prior anti-diabetic drugs, prior comorbidities, renal function, duration from T2DM diagnosis initial drug exposure, HbA1c and fasting glucose were performed using the nearest neighbour search strategy with a calliper of 0.1. Propensity score matching was performed using Stata software (Version 16.0). Baseline characteristics between patients with SGLT2I and DPP4I use before and after matching were compared with absolute standardized mean difference (SMD), with SMD<0.10 regarded as well-balanced between the two groups.

The cumulative incidence curves for the primary outcomes and secondary outcomes were constructed. Proportional Cox regression models were used to identify significant risk predictors of adverse study outcomes, with adjustment for demographics, comorbidities, number of prior hospitalisations, medication profile, renal function, glycaemic tests, and the duration of T2DM. The log-log plot was used to verify the proportionality assumption for the proportional Cox regression models. Subgroup analysis was conducted to confirm the association amongst patients with different clinical important predictors accounting to the diabetic and the metabolic profile, as well as the conditions and medications associated with prostate diseases. Cause-specific and sub-distribution hazard models were conducted to consider possible competing risks. Multiple propensity adjustment approaches were used, including propensity score stratification [20], propensity score matching with inverse probability of treatment weighting [21] and propensity score matching with stable inverse probability weighting [22]. The three arm sensitivity results involving glucagon-like peptide-1 receptor agonist (GLP1a) using stabilized IPTW were conducted to test the association and choice amongst the novel second-line anti-diabetic medications. Patients with CKD stage 4/5 (eGFR <30 mL/min/1.73m^2), peritoneal dialysis or haemodialysis who may be contraindicated with SGLT2I were excluded in the sensitivity analysis. Sensitivity analysis results with consideration of one-year lag time effects was also conducted. The hazard ratio (HR), 95% CI and P-value were reported. Statistical significance was defined as P-value <0.05. All statistical analyses were performed with RStudio (Version: 1.1.456) and Python (Version: 3.6), unless otherwise specified.

## Results

### Basic characteristics

In this territory-wide cohort study of 42491 male patients with T2DM treated with SGLT2I or DPP4I between 1st January 2015 and 31st December 2020 in Hong Kong, patients were followed up until 31st December 2020 or until their deaths (**Figure 1**). After exclusion, This cohort comprises 42129 male T2DM patients with a mean age of 61.0 years old (Standard deviation: 12.2), amongst which 17120 patients used SGLT2Is, and 25009 patients used DPP4Is.

The DPP4I and SGLT2I cohorts were comparable after matching with nearest neighbour search strategy with calliper of 0.1, and the proportional hazard assumption was confirmed **(Supplementary Figure 1**). After the propensity score matching, the baseline characteristic of the SGLT2I and the DPP4I users were well-balanced, apart from sulphonylurea (SMD=0.17) and thiozolidinedone (SMD=0.11) (**Table 1**). The 2 unbalanced variables were adjusted in the Cox regression models and were included in the subgroup analysis. In the matched cohort, 162 patients (0.47%) developed new onset prostate cancer. The characteristics of patients are shown in **Table 1**.

**Table 1.** Baseline and clinical characteristics of patients with SGLT2I v.s. DPP4I use before and after propensity score matching (1:1) * for SMD≥0.1; SGLT2I: sodium glucose cotransporter-2 inhibitor; DPP4I: dipeptidyl peptidase-4 inhibitor; MDRD: modification of diet in renal disease; ACEI: angiotensin-converting enzyme inhibitors; ARB: angiotensin II receptor blockers.

| Characteristics | Before matching |  |  |  | After matching |  |  |  |
| --- | --- | --- | --- | --- | --- | --- | --- | --- |
|  | All (N=42129)<br>Mean(SD);N or<br>Count(%) | SGLT2I (N=17120)<br>Mean(SD);N or<br>Count(%) | DPP4I (N=25009)<br>Mean(SD);N or<br>Count(%) | SMD | All (N=34240)<br>Mean(SD);N or<br>Count(%) | SGLT2I (N=17120)<br>Mean(SD);N or<br>Count(%) | DPP4I (N=17120)<br>Mean(SD);N or<br>Count(%) | SMD |
| **Demographics** |  |  |  |  |  |  |  |  |
| Baseline age, years | 61.0(12.2);n=42129 | 57.3(10.9);n=17120 | 63.5(12.4);n=25009 | 0.53* | 57.7(10.7);n=34240 | 57.3(10.9);n=17120 | 58.2(10.5);n=17120 | 0.08 |
| **Past comorbidities** |  |  |  |  |  |  |  |  |
| Charlson standard comorbidity index | 1.9(1.5);n=42129 | 1.5(1.2);n=17120 | 2.2(1.6);n=25009 | 0.47* | 1.5(1.2);n=34240 | 1.5(1.2);n=17120 | 1.6(1.2);n=17120 | 0.05 |
| Duration from earliest diabetes mellitus diagnosis date to baseline date, years | 6.5(4.9);n=42129 | 6.8(4.9);n=17120 | 6.3(4.9);n=25009 | 0.1 | 6.7(5.0);n=34240 | 6.8(4.9);n=17120 | 6.5(5.1);n=17120 | 0.07 |
| Number of prior hospitalizations | 1.2(0.8);n=42129 | 1.3(1.1);n=17120 | 1.2(0.6);n=25009 | 0.17 | 1.3(1.0);n=34240 | 1.3(1.1);n=17120 | 1.2(0.9);n=17120 | 0.08 |
| Hyperlipidaemia | 30441(72.25%) | 12812(74.83%) | 17629(70.49%) | 0.1 | 25853(75.50%) | 12812(74.83%) | 13041(76.17%) | 0.03 |
| Hypertension | 25490(60.50%) | 15280(89.25%) | 10210(40.82%) | 1.18* | 24440(71.37%) | 12280(71.72%) | 12160(71.02%) | 0.02 |
| Urinary tract infection | 1493(3.54%) | 415(2.42%) | 1078(4.31%) | 0.1 | 807(2.35%) | 415(2.42%) | 392(2.28%) | 0.01 |
| Cancer | 731(1.73%) | 209(1.22%) | 522(2.08%) | 0.07 | 415(1.21%) | 209(1.22%) | 206(1.20%) | <0.01 |
| Autoimmune disease | 330(0.78%) | 137(0.80%) | 193(0.77%) | <0.01 | 272(0.79%) | 137(0.80%) | 135(0.78%) | <0.01 |
| Heart failure | 1328(3.15%) | 495(2.89%) | 833(3.33%) | 0.03 | 979(2.85%) | 495(2.89%) | 484(2.82%) | <0.01 |
| Ischemic heart disease | 5200(12.34%) | 2677(15.63%) | 2523(10.08%) | 0.17 | 5013(14.64%) | 2577(15.05%) | 2436(14.22%) | 0.02 |
| Acute myocardial infarction | 1541(3.65%) | 771(4.50%) | 770(3.07%) | 0.07 | 1513(4.41%) | 771(4.50%) | 742(4.33%) | 0.01 |
| Peripheral vascular disease | 341(0.80%) | 98(0.57%) | 243(0.97%) | 0.05 | 196(0.57%) | 98(0.57%) | 98(0.57%) | <0.01 |
| Atrial fibrillation | 993(2.35%) | 370(2.16%) | 623(2.49%) | 0.02 | 724(2.11%) | 370(2.16%) | 354(2.06%) | 0.01 |
| Stroke/transient ischemic attack | 1259(2.98%) | 423(2.47%) | 836(3.34%) | 0.05 | 836(2.44%) | 423(2.47%) | 413(2.41%) | <0.01 |
| Diabetic retinopathy | 2723(6.46%) | 1019(5.95%) | 1704(6.81%) | 0.04 | 1926(5.62%) | 1019(5.95%) | 907(5.29%) | 0.03 |
| Diabetic nephropathy | 528(1.25%) | 116(0.67%) | 412(1.64%) | 0.09 | 232(0.67%) | 116(0.67%) | 116(0.67%) | <0.01 |
| Diabetic neuropathy | 872(2.06%) | 311(1.81%) | 561(2.24%) | 0.03 | 601(1.75%) | 311(1.81%) | 290(1.69%) | 0.01 |
| Renal diseases | 3269(7.75%) | 899(5.25%) | 2370(9.47%) | 0.16 | 1704(4.97%) | 899(5.25%) | 805(4.70%) | 0.03 |
| Liver diseases | 1741(4.13%) | 807(4.71%) | 934(3.73%) | 0.05 | 1520(4.43%) | 807(4.71%) | 713(4.16%) | 0.03 |
| Chronic obstructive pulmonary disease | 466(1.10%) | 122(0.71%) | 344(1.37%) | 0.07 | 243(0.70%) | 122(0.71%) | 121(0.70%) | <0.01 |
| **Medications** |  |  |  |  |  |  |  |  |
| Number of anti-diabetic drugs | 9.5(26.7);n=42129 | 12.7(32.8);n=17120 | 7.3(21.3);n=25009 | 0.2 | 12.7(32.8);n=34240 | 12.7(32.8);n=17120 | 12.8(32.7);n=17120 | <0.01 |
| Metformin | 37650(89.36%) | 15922(93.00%) | 21728(86.88%) | 0.20* | 31909(93.19%) | 15922(93.00%) | 15987(93.38%) | 0.02 |
| Sulphonylurea | 32456(77.03%) | 12393(72.38%) | 20063(80.22%) | 0.19 | 26040(76.05%) | 12393(72.38%) | 13647(79.71%) | 0.17* |
| Insulin | 21040(49.94%) | 8455(49.38%) | 12585(50.32%) | 0.02 | 16773(48.98%) | 8455(49.38%) | 8318(48.58%) | 0.02 |
| Acarbose | 1402(3.32%) | 640(3.73%) | 762(3.04%) | 0.04 | 1235(3.60%) | 640(3.73%) | 595(3.47%) | 0.01 |
| Thiazolidinedione | 8506(20.19%) | 4448(25.98%) | 4058(16.22%) | 0.24* | 8131(23.74%) | 4448(25.98%) | 3683(21.51%) | 0.11* |
| Glucagon-like peptide-1 receptor agonists | 2666(6.32%) | 1159(6.76%) | 1507(6.02%) | 0.03 | 2250(6.57%) | 1159(6.76%) | 1091(6.37%) | 0.02 |
| Statins and fibrates | 26683(63.33%) | 12716(74.27%) | 13967(55.84%) | 0.39* | 25225(73.67%) | 12716(74.27%) | 12509(73.06%) | 0.03 |
| ACEI/ARB | 22307(52.94%) | 10563(61.69%) | 11744(46.95%) | 0.30* | 21120(61.68%) | 10563(61.69%) | 10557(61.66%) | <0.01 |
| Diuretics | 12276(29.13%) | 5536(32.33%) | 6740(26.95%) | 0.12 | 11411(33.32%) | 5536(32.33%) | 5875(34.31%) | 0.04 |
| Beta-blockers | 11878(28.19%) | 5048(29.48%) | 6830(27.31%) | 0.05 | 10565(30.85%) | 5048(29.48%) | 5517(32.22%) | 0.06 |
| Calcium channel blockers (dihydropyridine) | 21531(51.10%) | 9919(57.93%) | 11612(46.43%) | 0.23* | 19921(58.18%) | 9919(57.93%) | 10002(58.42%) | 0.01 |
| Calcium channel blocker (non-dihydropyridine) | 11656(27.66%) | 5014(29.28%) | 6642(26.55%) | 0.06 | 9795(28.60%) | 5014(29.28%) | 4781(27.92%) | 0.03 |
| Systemic steroids | 269(0.63%) | 126(0.73%) | 143(0.57%) | 0.02 | 252(0.73%) | 126(0.73%) | 126(0.73%) | <0.01 |
| Antiplatelets | 13825(32.81%) | 6401(37.38%) | 7424(29.68%) | 0.16 | 12869(37.58%) | 6401(37.38%) | 6468(37.78%) | 0.01 |
| Alpha-blockers | 7488(17.77%) | 2624(15.32%) | 4864(19.44%) | 0.11 | 5033(14.69%) | 2624(15.32%) | 2409(14.07%) | 0.04 |
| Aspirin | 12408(29.45%) | 5963(34.83%) | 6445(25.77%) | 0.2 | 11902(34.76%) | 5963(34.83%) | 5939(34.69%) | <0.01 |
| **Calculated biomarker** |  |  |  |  |  |  |  |  |
| Abbreviated MDRD, mL/min/1.73m<sup>2</sup> | 80.8(26.9);n=34221 | 88.6(22.6);n=14077 | 75.4(28.3);n=20144 | 0.51* | 87.5(23.2);n=28366 | 88.6(22.6);n=14077 | 86.4(23.7);n=14289 | 0.09 |
| **Renal function tests** |  |  |  |  |  |  |  |  |
| Creatinine, umol/L | 103.7(79.5);n=34221 | 87.9(29.0);n=14077 | 114.8(99.2);n=20144 | 0.37* | 89.7(37.2);n=28366 | 87.9(29.0);n=14077 | 91.4(43.8);n=14289 | 0.09 |
| **Lipid tests** |  |  |  |  |  |  |  |  |
| Triglyceride, mmol/L | 1.7(1.8);n=32353 | 1.9(2.0);n=13493 | 1.7(1.6);n=18860 | 0.1 | 1.9(2.0);n=27134 | 1.85(1.96);n=13493 | 1.87(2.08);n=13641 | 0.01 |
| Low-density lipoprotein, mmol/L | 2.4(0.8);n=31726 | 2.3(0.8);n=13219 | 2.4(0.8);n=18507 | 0.04 | 2.4(0.8);n=26546 | 2.3(0.8);n=13219 | 2.4(0.8);n=13327 | 0.05 |
| High-density lipoprotein, mmol/L | 1.1(0.3);n=32298 | 1.09(0.28);n=13468 | 1.14(0.31);n=18830 | 0.14 | 1.1(0.3);n=27093 | 1.09(0.28);n=13468 | 1.1(0.29);n=13625 | 0.04 |
| Total cholesterol, mmol/L | 4.2(1.0);n=32387 | 4.22(1.01);n=13510 | 4.23(0.99);n=18877 | 0.01 | 4.3(1.0);n=27166 | 4.2(1.0);n=13510 | 4.3(1.0);n=13656 | 0.06 |
| **Glucose tests** |  |  |  |  |  |  |  |  |
| Haemoglobin A1C, % | 8.1(1.6);n=33538 | 8.3(1.6);n=13824 | 8.0(1.6);n=19714 | 0.18 | 8.3(1.6);n=27789 | 8.25(1.61);n=13824 | 8.26(1.58);n=13965 | 0.01 |
| Fasting glucose, mmol/L | 9.0(3.9);n=30477 | 9.2(3.6);n=12691 | 8.8(4.2);n=17786 | 0.08 | 9.2(3.8);n=25612 | 9.2(3.6);n=12691 | 9.3(4.1);n=12921 | 0.04 |

### Primary analysis

In the matched cohort, 60 SGLT2I users and 102 DPP4I users developed prostate cancer. In a total of 186869.4 person-year after a follow-up of a median of 5.61 years, the incidence of prostate cancer was lower amongst SGLT2I users (Incidence rate, IR: 0.63; 95% confidence interval, CI: 0. 48-0.81) than in DPP4I (IR: 1.11; CI: 0.90-1.34) users. SGLT2 inhibitors users had a 55% lower risk of prostate cancer compared to users of DPP-4 inhibitors after adjustment for demographics, comorbidities, number of prior hospitalisations, medication profile, renal function, glycaemic tests, and the duration of T2DM (HR: 0.45; 95% CI: 0.30-0.70, p=0.0003) (**Table 2; Figure 2; Supplementary Table 2)**. This was substantiated by the cumulative incidence curves stratified by SGLT2I versus DPP4I (**Figure 2**). The marginal effects plots demonstrated that SGLT2I was associated with lower risks of prostate cancer regardless of the duration of the diabetes diagnosis (**Supplementary Figure 2**), regardless of the number of prior hospitalizations (**Supplementary Figure 3**).

**Figure 2.**
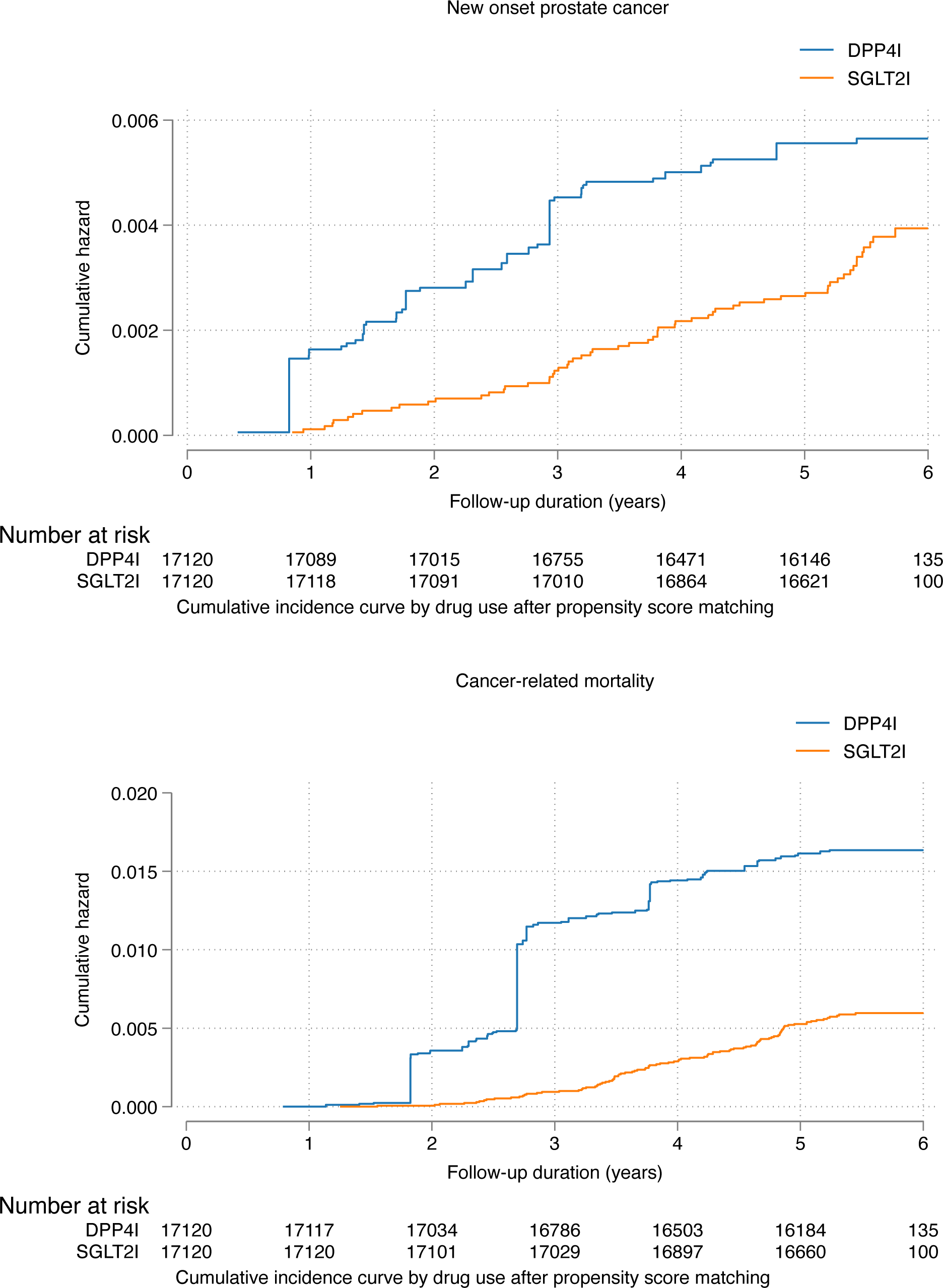
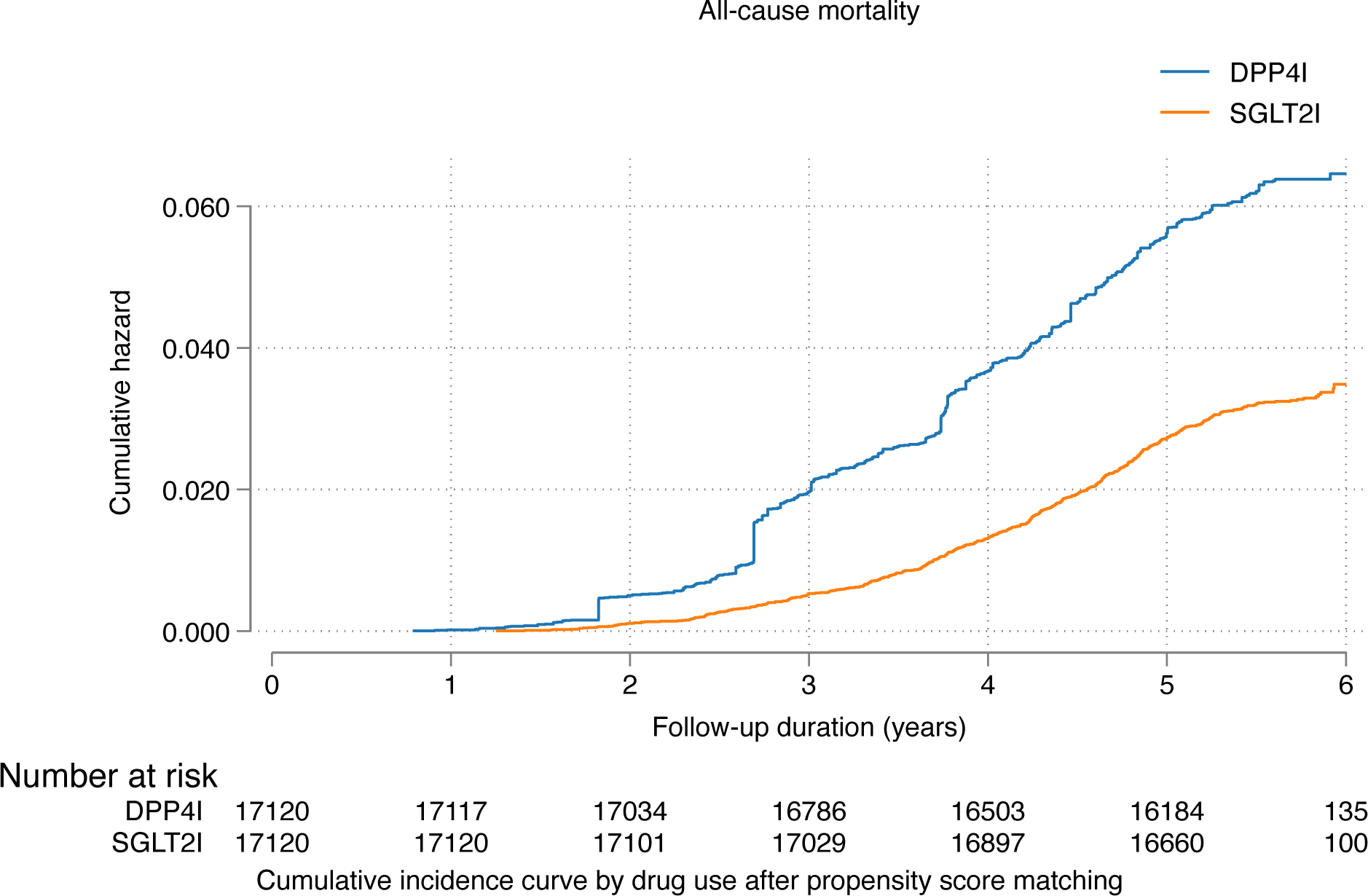
Cumulative incidence curves for new onset prostate cancer, cancer-related mortality, and all-cause mortality stratified by drug exposure effects of SGLT2I and DPP4I after propensity score matching (1:1) SGLT2I: Sodium-glucose cotransporter-2 inhibitors; DPP4I: Dipeptidyl peptidase-4 inhibitors

**Table 2.** Incidence rate (IR) per 1000 person-year of new onset prostate cancer, cancer-related mortality, and all-cause mortality in the cohort before and after 1:1 propensity score matching. SGLT2I: sodium glucose cotransporter-2 inhibitor; DPP4I: dipeptidyl peptidase-4 inhibitor; MDRD: modification of diet in renal disease Adjusted for significant demographics, past comorbidities, duration of diabetes mellitus, number of prior hospitalizations, number of anti-diabetic drugs, non-SGLT2I/DPP4I medications, abbreviated MDRD, HbA1c, fasting glucose.

| **Prostate cancer** | **Number of patients** | **Number of events** | **Total person-year** | **Follow-up duration, years [95% CI]** | **Incidence [95% CI]** | **Adjusted hazard ratio [95% CI]** | **P-value** |
| --- | --- | --- | --- | --- | --- | --- | --- |
| **DPP4I** | 17120 | 102 | 92074.6 | 5.6 [5.31-5.83] | 1.11[0.90-1.34] | 1 [reference] | NA |
| **SGLT2I** | 17120 | 60 | 94794.8 | 5.62 [5.35-5.83] | 0.63[0.48-0.81] | 0.45[0.30-0.70] | 0.0003 |
| **Cancer-related mortality** | **Number of patients** | **Number of events** | **Total person-year** | **Follow-up duration, years [95% CI]** | **Incidence [95% CI]** | **Adjusted hazard ratio [95% CI]** | **P value** |
| **DPP4I** | 17120 | 260 | 92299.7 | 5.6 [5.31-5.83] | 2.81[2.48-3.18] | 1 [reference] | NA |
| **SGLT2I** | 17120 | 99 | 94892.2 | 5.62 [5.33-5.82] | 1.04[0.85-1.27] | 0.57[0.40-0.76] | <0.0001 |
| **All-cause mortality** | **Number of patients** | **Number of events** | **Total person-year** | **Follow-up duration, years [95% CI]** | **Incidence [95% CI]** | **Adjusted hazard ratio [95% CI]** | **P value** |
| **DPP4I** | 17120 | 1652 | 92299.7 | 5.6 [5.31-5.83] | 17.9[17.0-18.8] | 1 [reference] | NA |
| **SGLT2I** | 17120 | 541 | 94892.2 | 5.62 [5.35-5.82] | 5.70[5.23-6.20] | 0.68[0.46-0.78] | <0.0001 |

In a total of 187191.9 person-year, 260 DPP4I and 99 SGLT2I users passed away due to cancer. The incidence of cancer-related mortality was lower amongst SGLT2I users (IR: 1.04; 95% CI: 0.85-1.27) compared to DPP4I users (IR: 2.81; 95% CI: 2.48-3.18). SGLT2I users had a 43% lower risk of cancer-related mortality after adjustment (HR: 0.55; 95% CI: 0.50-0.61, p<0.0001) compared to users of DPP4I. Besides, 1652 DPP4I users and 541 SGLT2I users passed away. SGLT2I users had a 32% lower risk of all-cause mortality after adjustment (HR: 0.68; 95% CI: 0.46-0.78, p<0.0001) compared to users of DPP4I (**Table 2; Figure 2; Supplementary Table 2**).

### Subgroup analysis and sensitivity analysis

The results of the subgroup analysis for the effects of SGLT2I and DPP4I on prostate cancer are shown in **Figure 3**. In the subgroup analysis, the results demonstrated that SGLT2I was associated with lower risks of prostate cancer amongst patients who were older (age >65). The results were consistent amongst patients with quartile 2 and 3 HbA1c. Besides, SGLT2I was associated with lower risks of prostate cancer amongst patients without prior heart failure, acute myocardial infarction, atrial fibrillation, peripheral vascular disease, cancer, and renal disease (all p<0.05). Furthermore, SGLT2I had lower risks of prostate cancer amongst patients without using alpha-blockers and antiplatelet (all p<0.05). SGLT2I was also associated with lower risks of prostate cancers amongst patients not on insulin, thiazolidinedione, GLP1a. However, those could be due to the insufficient sample size in the subgroups amongst the patients with the diseases or drug users. SGLT2I was also associated with lower risks of prostate cancer amongst metformin and sulphonylurea users. Indeed, SGLT2I was associated with higher risks of prostate cancer amongst patients without sulphonylurea (HR: 5.55; 95% CI: 1.26-22.43).

**Figure 3.**
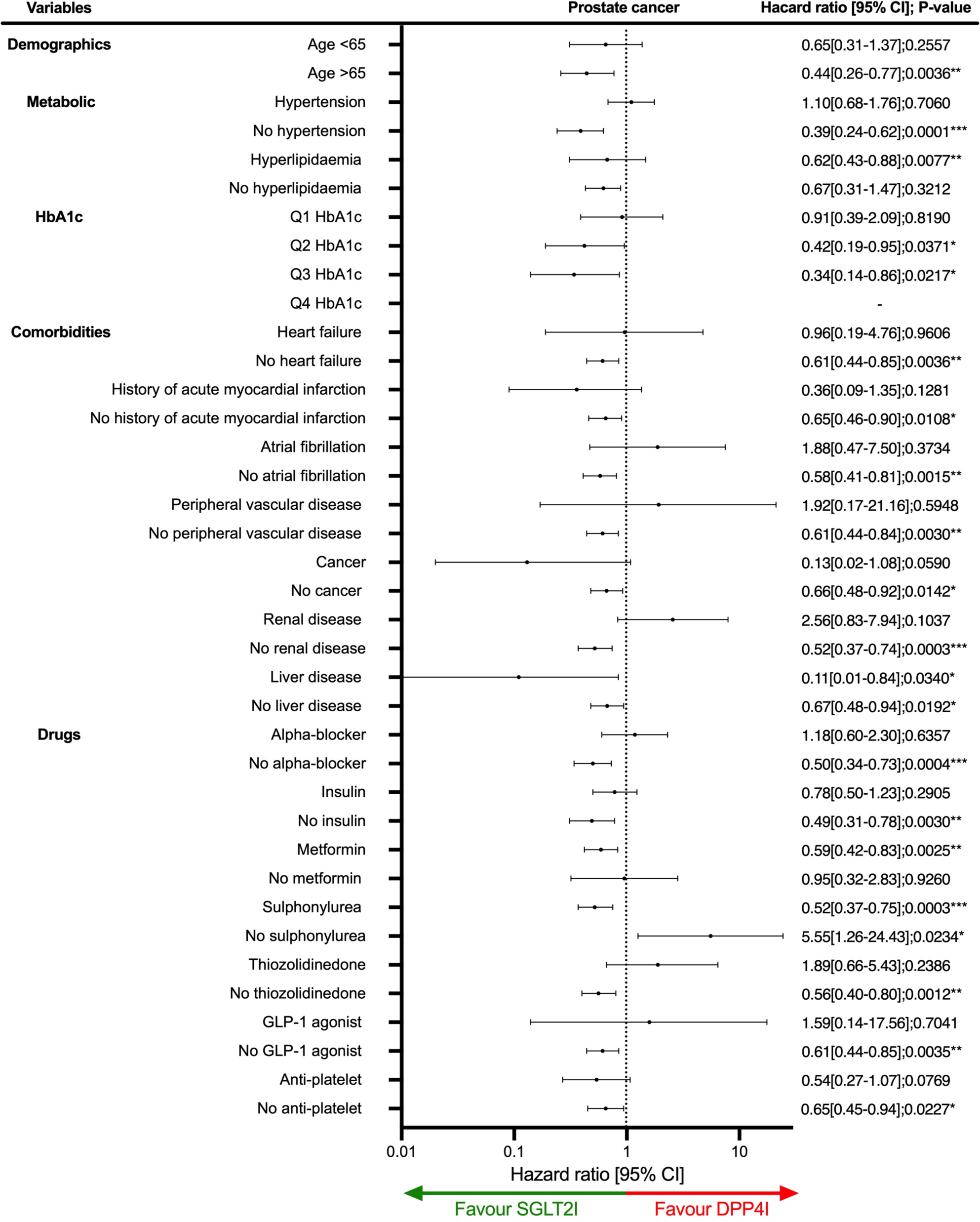
Subgroup analyses for SGLT2I v.s. DPP4I exposure predict prostate cancer in the matched cohort. SGLT2I: Sodium-glucose cotransporter-2 inhibitors; DPP4I: Dipeptidyl peptidase-4 inhibitors.Q1: Quartile 1Q2: Quartile 2; Q3: Quartile 3; Q4: Quartile 4; CI: Confidence interval

### Sensitivity analysis

Sensitivity analyses were performed to confirm the predictability of the models. The results of the cause-specific hazard models, sub-distribution hazard models and different propensity score approaches demonstrated that different models do not change the point estimates for both the primary and the secondary outcomes (all P<0.05) **(Supplementary Table 3).** A 3-arm analysis with the inclusion of GLP1a including patients only on SGLT2I, DPP4I, and GLP1a using stabilized IPTW was conducted (**Supplementary Table 4**). The results between DPP4I and SGLT2I remained consistent with the main result (all p<0.05) (**Table 2**). However, it was found that SGLT2I was not associated with lower risks of prostate cancer compared to GLP1a. Excluding patients with CKD stage 4/5 (eGFR <30 mL/min/1.73m^2), peritoneal dialysis or haemodialysis in the matched cohort likewise demonstrated that SGLT2I was associated with lower risks of prostate cancer compared to DPP4I **(Supplementary Table 5)**. Meanwhile, the sensitivity analysis for 1-year lag time also demonstrated the same trend (all p<0.05) **(Supplementary Table 6)**.

## Discussion

In this territory-wide cohort study, we utilised real-world data to compare the association between SGLT2I, DPP4I and new-onset prostate cancer. Our results suggest that SGLT2I usage is associated with a lowered risk of new-onset prostate cancer than DPP4I usage. To the best of our knowledge, this is the first study to directly compare the relative association of two anti-diabetic medications on the incidence of prostate cancer.

### Comparison with previous studies

The prevalence of prostate cancer in our study (IR: 86.7; 95% CI: 73.9-101 per 100,000 person-year) is like the existing literature, which described an incidence of 65.1 per 100,000 person-year amongst T2DM patients in the whole China [23–25]. It was believed that T2DM patients are associated with an increased prevalence of prostate cancer and adverse complications [26]. A meta-analysis suggests that diabetic patients are associated with a worse prognosis [27]. On the contrary, some studies show no correlation or even an inverse association between T2DM and prostate cancer [28]. While the relationship between T2DM and prostate cancer remained controversial, the pleiotropic effects of the anti-diabetic drugs may provide us with new insights.

In our study, we found that SGLT2I users had 55% relatively lower risks of prostate cancer compared to DPP4I users. In a previous meta-analysis, it was demonstrated that there was no association between SGLT2I use and incidence of overall cancer [29]. A meta-analysis found that DPP4I and SGLT2I show no significant association with prostate cancer compared to placebo, while GLP1a agents demonstrated beneficial effects against prostate cancer [30]. However, the 7 randomised control trials studies included regarding of SGLT2I were posthoc analyses without the primary aim of investigating prostate cancer, and the results demonstrated a relatively high heterogenicity (P_heterogenity_<0.01, *I^2^* = 97.3%). The meta-analysis did not directly compare the effects between SGLT2I and DPP4I. For the SGLT2I-DPP4I comparison, Rokszin *et al.* found no significant difference in the risk of prostate cancer (HR: 0.65; 95% CI: 0.42-1.02) between the SGLT2I and DPP4I users [12]. However, the difference between the two arms remained worthy of investigation since the upper 95% CI of prostate cancer was close to 1 and that study followed up the patients for two years only, while in our study the follow-up duration median was 5.61 years.

The investigation of novel anti-diabetic medications was prompted by biological evidence in the current literature that underscores possible pleiotropic effects of SGLT2I and DPP4I on prostate cancer [31–33]. Interest in this area was prompted largely due to the fact that cancer cells utilise glucose as a primary source of fuel rather than fatty acids [34]. Moreover, evidence suggests that SGLT2I usage may inhibit the AMPK/mTOR pathway and induce apoptosis of cancerous cells [35]. Scafoglio *et al.* found abundant functional expression of SGLT2 on prostate adenocarcinomas; thereby inhibition of SGLT2 receptors reduces glucose uptake and tumour progression in a xenograft model [36]. In a cell culture study, it was found that SGLT2I inhibited the clonogenic survival of prostate cancer cells [37]. In our study, the protective effects of SGLT2I were significant amongst patients without comorbidities but not those with prior comorbidities (heart failure, prior acute myocardial infarction, atrial fibrillation, peripheral vascular diseases). On one hand, while this could be explained by the limited sample size in the subgroups, we may also hypothesise that this could be explained partially by the SGLT2 expression level, as systemic diseases were suggested to increase the expression and activity of SGLT2 [38].

It must be noted that this does not necessarily mean that DPP4I is harmful in terms of new-onset prostate cancer, but it may as well be protective against prostate cancer compared to non-users. DPP4I and SGLT2I may modulate the risks of developing prostate cancer amongst T2DM patients. For DPP4I, previously, it was found that the use of saxagliptin and exenatide may decrease prostate cancer risk compared to sulfonylurea usage [39]. In another observational study, sitagliptin usage was associated with lower risks of prostate cancer amongst Asians with an odd ratio of 0.61 [40]. In addition, the Liraglutide Effect and Action in Diabetes: Evaluation of Cardiovascular Outcome Results (LEADER) trial revealed that malignant prostate neoplasms were present in a lower proportion of diabetic patients who use liraglutide compared to the placebo group [41]. Although the mechanism behind the observed effects of cancer growth inhibition remains ambiguous, the potential implications of SGLT2I and DPP4I usage as a part of cancer treatment continue to be revised in the growing body of literature.

### Clinical implications

The potential antineoplastic benefit of SGLT2I and DPP4I on various cancer types have received global interest in recent years. As prostate cancer is one of the leading causes of cancer-related mortality among men, our findings could carry critical clinical implications in optimising treatment for T2DM male patients who might be at higher risk of developing prostate cancer. Thus, future studies in-cooperating randomised clinical trials are essential for clinicians to provide prevention of prostate cancer for diabetic patients. In the long term, this can reduce healthcare resource expenditure and refine clinical guidelines that govern treatment for prostate cancer and diabetes. This study has explored the relationship of data obtained from routine clinical practice in Hong Kong. This study suggests that SGLT2I may demonstrate a more significant overall protective effect against prostate cancer compared to DPP4I. Previously, metformin was suggested to be associated with reduced mortality amongst prostate cancer patients independent of diabetic control [42]. As such, SGLT2I may also play a role in altering the cancer outcomes and prognosis amongst prostate cancer patients.

### Limitations

Several limitations should be noted in this study. Due to the observational nature of this study variables which predict prostate cancer, such as BMI, alcohol use, smoking, socioeconomic status, family history of prostate cancer, and screening PSA level, were not available from CDARS. This was attempted to be addressed by including laboratory results and comorbidities to infer the potential risk factors. To further compensate for this, we have matched our drug cohorts over an array of diseases and medications, adjusted the regression, and conducted a range of sensitivity analyses to minimise bias. Besides, there is potential under-coding, coding errors, and missing data leading to information bias due to the nature of the study. Moreover, the patient’s level of compliance and drug exposure duration could not be standardised. Lastly, the retrospective design of our study necessitates the presentation of associations but not causal links between SGLT2I versus DPP4I use and the risk of new-onset prostate cancer. Therefore, further research is warranted to explore the beneficial effects of SGLT2I.

## Conclusion

In this population-based cohort study, SGLT2I users were associated with lower risks of new-onset prostate cancer than DPP4I users amongst T2DM patients. These results may have potential clinical implications in preventing cancer among T2DM patients. Further randomised controlled trials and mechanistic studies into prostate cancer with SGLT2I are needed.

## Ethical approval statement

This study was approved by the Institutional Review Board of the University of Hong Kong/Hospital Authority Hong Kong West Cluster (HKU/HA HKWC IRB) (UW-20-250), and New Territories East Cluster-Chinese University of Hong Kong (NTEC-UCHK) Clinical Research Ethnics Committee (2018.309, 2018.643) and complied with the Declaration of Helsinki.

## Availability of data and materials

Data are not available, as the data custodians (the Hospital Authority and the Department of Health of Hong Kong SAR) have not given permission for sharing due to patient confidentiality and privacy concerns. Local academic institutions, government departments, or nongovernmental organizations may apply for the access to data through the Hospital Authority’s data sharing portal (https://www3.ha.org.hk/data).

## Supporting information

STROBE_checklist

## Acknowledgements

None.

## Guarantor Statement

All authors approved the final version of the manuscript. GT is the guarantor of this work and, as such, had full access to all the data in the study and takes responsibility for the integrity of the data and the accuracy of the data analysis.

## Author contributions

Data analysis: OHIC, LL, JZ

Data review: OHIC, JSKC, RNCC, SL, GT, JZ

Data acquisition: OHIC, HHHP, SL

Data interpretation: OHIC, CTC, JSKC, BMYC, GT, JZ

Critical revision of manuscription: JSKC, RNCC, ALYH, ECD, KN, BMYC, GT, JZ Supervision: BMYC, GT, JZ

Manuscript writing: OHIC, CTC, JSKC, ALYH, KKW

Manuscript revision: OHIC, JSKC, ALYH, ECD, KN, BMYC, GT, JZ

