## Supplementary Appendix for "New-onset prostate cancer in type 2 diabetes mellitus exposed to the SGLT2I, DPP4I and GLP1a: A population-based cohort study"

**Table of Contents**

*Supplementary Figure 1. Propensity score matching comparisons and proportional hazard assumption checking with parallel lines for SGLT2I versus DPP4I before and after 1:1 matching with nearest neighbours search strategy with calliper of 0.1* ........................... 2

*Supplementary Figure 2. Marginal effects of previous diabetes duration with 95% CIs on new onset prostate cancer, cancer-related mortality, and all-cause mortality stratified by drug use in the matched cohort.* ........................................................................................ 4

*Supplementary Figure 3. Marginal effects of number of prior hospitalizations with 95% Cis on new onset prostate cancer, cancer-related mortality, and all-cause mortality stratified by drug use in the matched cohort.* ................................................ Error! Bookmark not defined.

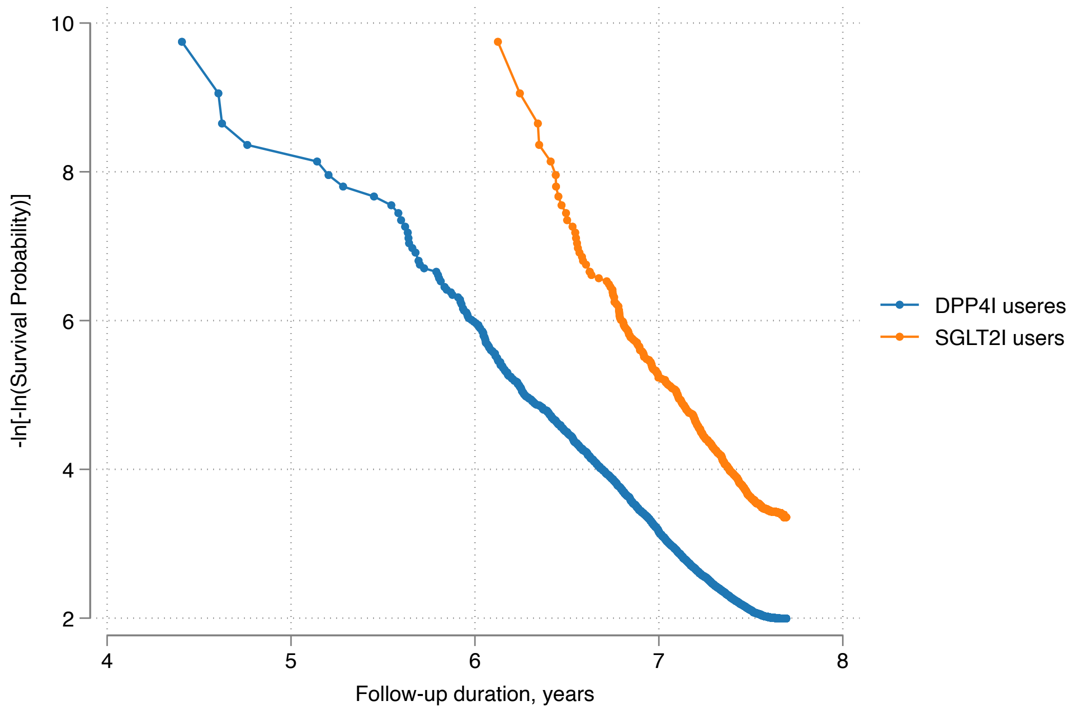

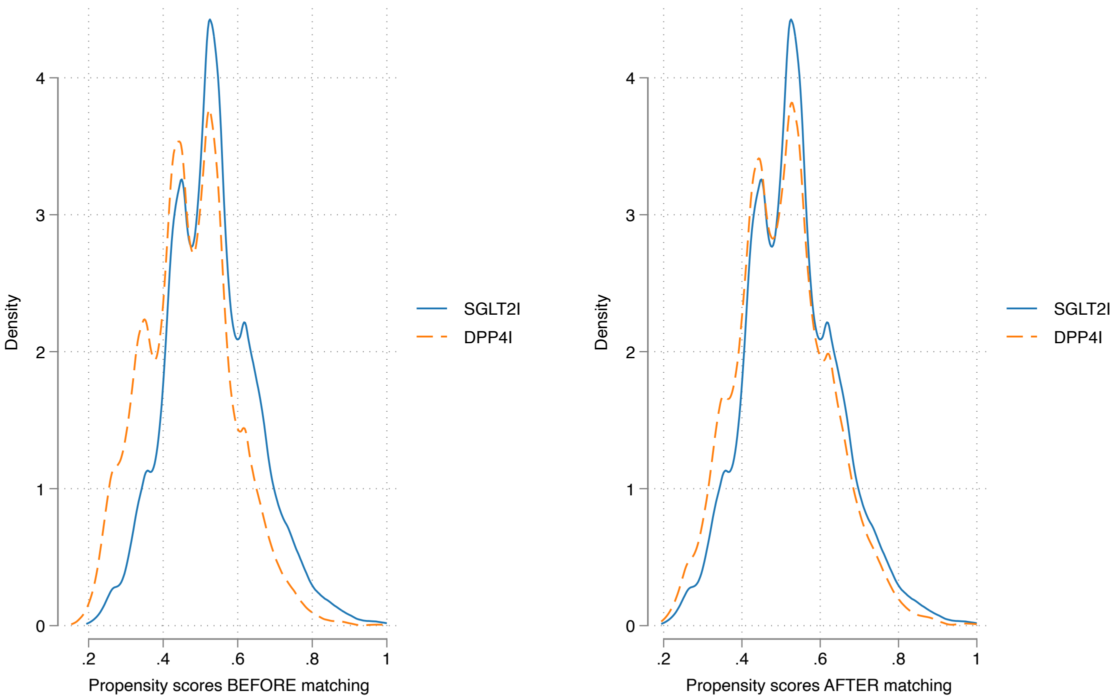

Nearest neighbor search strategy with caliper=0.1.

**Supplementary Figure 1. Propensity score matching comparisons and proportional hazard assumption checking with parallel lines for SGLT2I versus DPP4I before and after 1:1 matching with nearest neighbours search strategy with calliper of 0.1**

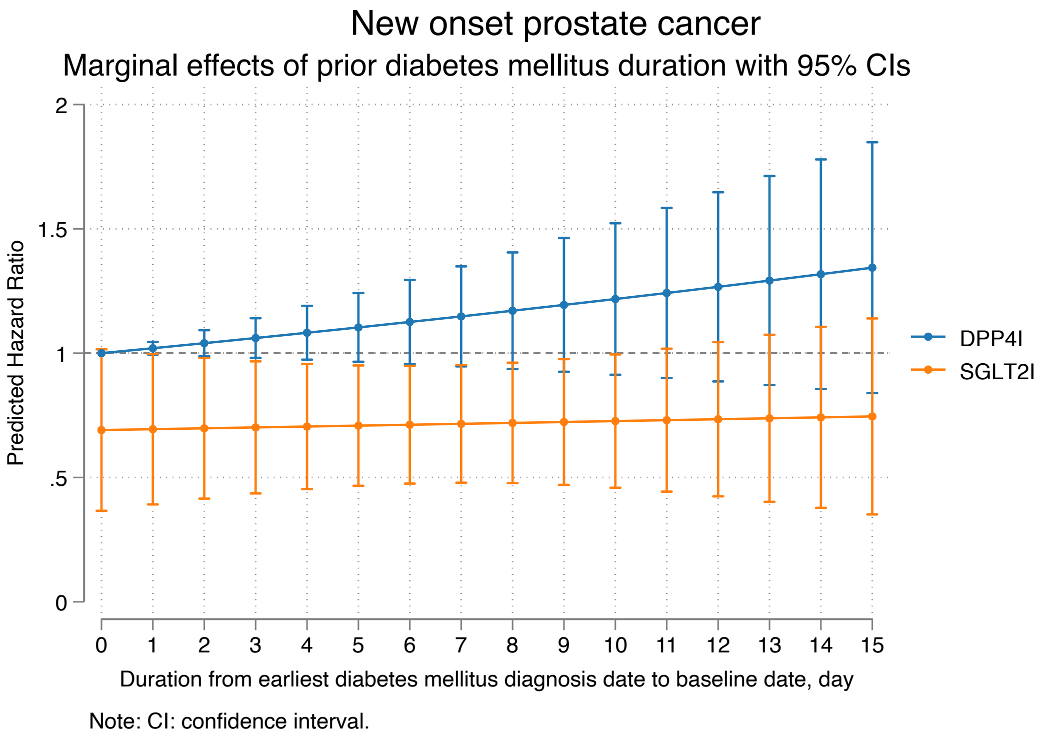
New onset prostate cancer
Marginal effects of prior diabetes mellitus duration with 95% CIs
Predicted Hazard Ratio
0
.5
1
1.5
2
0 1 2 3 4 5 6 7 8 9 10 11 12 13 14 15
Duration from earliest diabetes mellitus diagnosis date to baseline date, day
DPP4I
SGLT2I
Note: CI: confidence interval.

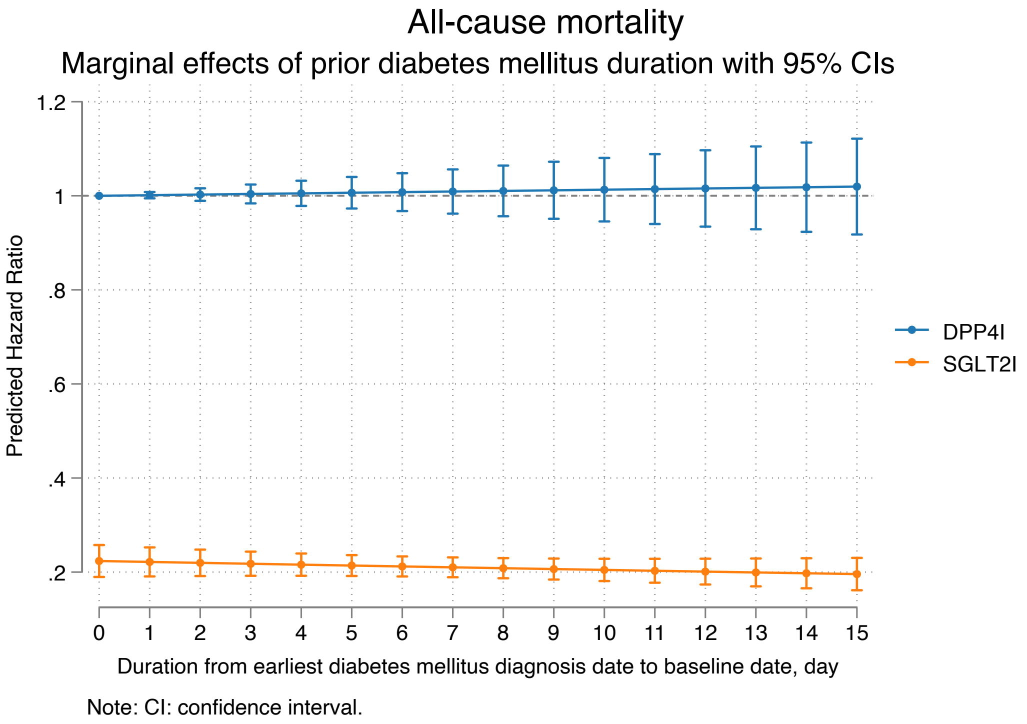
All-cause mortality
Marginal effects of prior diabetes mellitus duration with 95% CIs
Predicted Hazard Ratio
.2
.4
.6
.8
1
1.2
0 1 2 3 4 5 6 7 8 9 10 11 12 13 14 15
Duration from earliest diabetes mellitus diagnosis date to baseline date, day
DPP4I
SGLT2I
Note: CI: confidence interval.

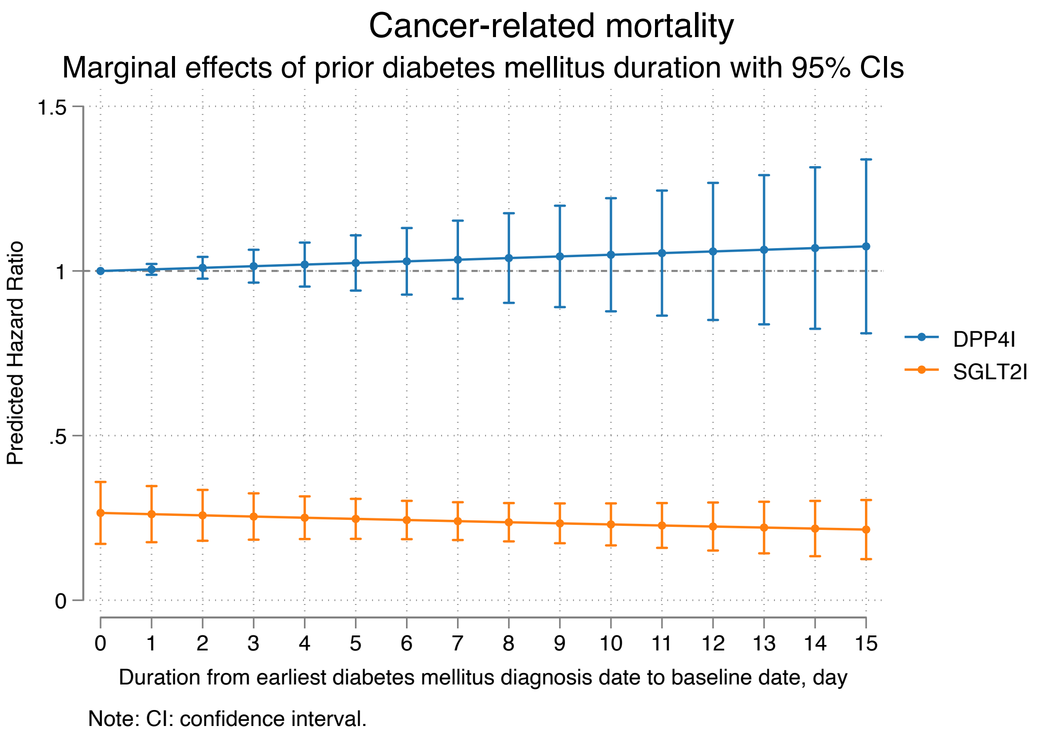

**Supplementary Figure 2. Marginal effects of previous diabetes duration with 95% CIs on new onset prostate cancer, cancer-related mortality, and all-cause mortality stratified by drug use in the matched cohort.**

SGLT2I: Sodium-glucose cotransporter-2 inhibitors; DPP4I: Dipeptidyl peptidase-4 inhibitors.

**Supplementary Table 1. The International Classification of Diseases, Clinical Modification (ICD-9-CM) codes for definitions of past comorbidities and outcomes.**

| Adverse outcome of interest |
|---|
| **Prostate cancer:** 185 |
| **Past comorbidities** |
| **Cancer**: 140-239 exclude prostate cancer |
| **Hypertension**: 401 401.1 401.9 402 402.01 402.1 402.11 402.9 402.91 403 403.01 403.1 403.11 403.9 403.91 404 404.01 404.02 404.03 404.1 404.11 404.12 404.13 404.9 404.91 404.92 404.93 405 405.01 405.09 405.1 405.11 405.19 405.9 405.91 405.99 437.2 + history of uses of anti-hypertensives |
| **Hyperlipidaemia**: 272.0 272.1 272.2 272.3 272.4 + history of uses of lipid-lowering drugs |
| **Liver disease**: 275.1 275.0 571.0 571.1 571.2 571.3 571.40 571.41 571.42 471.49 571.5 571.6 571.8, 571.9 572.0 572.4 572.1 572.3 572.8 573.0 573.4 573.8 573.9 |
| **Autoimmune diseases:** 136.1, 359.79, 359.71, 443.1, 446, 555, 556.0-556.6, 556.8-556.9, 695.4, 710, 714, 720, 725 |
| **Heart failure:** 428 428 428.1 428.2 428.2 428.21 428.22 428.23 428.3 428.3 428.31 428.32 428.33 428.4 428.4 428.41 428.42 428.43 428.9 398.91 402.01 402.11 402.91 404.01 404.03 404.11 404.13 404.91 404.93 |
| **Atrial fibrillation:** 427.31 429.4 |
| **Stroke/transient ischemic attack:** 435 435.1 435.2 435.3 435.8 435.9 433.81 433.91 434 436 437 437.1 433.31 433.01 434.01 434.1 434.11 434.9 434.91 437.2 437.3 437.4 437.5 437.6 437.7 437.8 437.9 430 431 432 432.1 432.9 |
| **Ischemic heart disease:** 410.01 410.02 410.1 410.11 410.12 410.2 410.21 410.22 410.3 410.31 410.32 410.4 410.41 410.42 410.5 410.51 410.52 410.6 410.61 410.62 410.7 410.71 410.72 410.8 410.81 410.82 410.9 410.91 410.92 411 411.1 411.8 411.81 411.89 413 413.1 413.9 414 414.01 414.02 414.03 414.04 414.05 414.06 414.07 414.1 414.11 414.12 414.19 414.2 414.3 414.4 414.8 414.9 410 412 |
| **Peripheral vascular disease:** 250.7 443.9 443 443.1 443.2 443.21 443.22 443.23 443.24 443.29 443.8 443.81 443.82 443.89 441 443.9 785.4 V43.4 |
| **Acute myocardial infarction:** 410 410.01 410.02 410.1 410.11 410.12 410.2 410.21 410.22 410.3 410.31 410.32 410.4 410.41 410.42 410.5 410.51 410.52 410.6 410.61 410.62 410.7 410.71 410.72 410.8 410.81 410.82 410.9 410.91 410.92 |
| **Renal diseases:** 39.95 54.98 403.01 403.11 403.91 404.02 404.03 404.12 404.13 404.92 404.91 582 582 582.1 582.2 582.4 582.8 582.81 582.89 582.9 583 583 583.1 583.2 583.4 583.6 583.7 584.x 585 585.1 585.2 585.3 585.4 585.5 585.6 585.9 586.x 588 588.1 588.8 588.81 588.89 588.9 + MDRD <60 |
| **Diabetic retinopathy:** 250.5 361 362.01 362.02 362.1 362.53 362.81 362.82 362.83 369 379.23 |
| **Diabetic nephropathy:** 250.40 250.41 250.42 250.43 |
| **Diabetic neuropathy**: 250.6, 337.0, 337.1, 354.0 – 355.9, 356.9, 357.2, 358.1, 536.3, 564.5, 596.54, 713.5, 951.0, 951.1, 951.3 |
| **Chronic obstructive pulmonary disease** 491.0 491,1 491.2 491.8 491.9 492.0 492.8 496 |

**Urinary tract infection:** 590, 595, 597, 590.1, 590.2, 590.8, 599.0, 771.82 + Urine culture results (*Escherichia* OR *Staphylococcus saprophyticus* OR *Klebsiella pneumoniae* OR *Proteus mirabilis* OR *Serratia marcescens* OR *Enterococci spp.* OR *Pseudomonas aeruginosa* OR *Enterobacter spp.* OR *Ureaplasma urealyticum* OR *Enterococcus spp.* OR *Citrobacter spp.* OR *Actinobacter spp.* OR *Streptococcus spp*)

**Supplementary Table 2. Multivariate Cox regression models with adjustments to predict new onset prostate cancer, cancer-related mortality, and all-cause mortality in the matched cohort.**
* for p≤ 0.05, ** for p ≤ 0.01, *** for p ≤ 0.001; HR: hazard ratio; CI: confidence interval; SGLT2I: sodium glucose cotransporter-2 inhibitor; DPP4I: dipeptidyl peptidase-4 inhibitor.
Model 1 adjusted for significant demographics.
Model 2 adjusted for significant demographics, and past comorbidities.
Model 3 adjusted for significant demographics, past comorbidities, duration from earliest diabetes mellitus date to initial drug exposure date, and number of prior hospitalizations.
Model 4 adjusted for significant demographics, past comorbidities, duration of diabetes mellitus, and number of prior hospitalizations, number of anti-diabetic drugs, and non-SGLT2I/DPP4I medications.
Model 5 adjusted for significant demographics, past comorbidities, duration of diabetes mellitus, and number of prior hospitalizations, number of anti-diabetic drugs, non-SGLT2I/DPP4I medications, abbreviated MDRD.
Model 6 adjusted for significant demographics, past comorbidities, duration of diabetes mellitus, and number of prior hospitalizations, number of anti-diabetic drugs, non-SGLT2I/DPP4I medications, abbreviated MDRD, HbA1c, fasting glucose.

| **Characteristics** | **New onset prostate cancer HR [95% CI];P value** | **Cancer-related mortality HR [95% CI];P value** | **All-cause mortality HR [95% CI];P value** |
|---|---|---|---|
| ***Model 1*** | 0.22[0.10-0.47];0.0001*** | 0.36[0.28-0.45];<0.0001*** | 0.51[0.46-0.57];<0.0001*** |
| ***Model 2*** | 0.29[0.12-0.70];0.0055** | 0.36[0.29-0.46];<0.0001*** | 0.53[0.48-0.59];<0.0001*** |
| ***Model 3*** | 0.30[0.16-0.56];0.0001*** | 0.37[0.29-0.46];<0.0001*** | 0.53[0.47-0.58];<0.0001*** |
| ***Model 4*** | 0.32[0.22-0.72];0.0051** | 0.37[0.29-0.46];<0.0001*** | 0.56[0.51-0.63];<0.0001*** |
| ***Model 5*** | 0.42[0.27-0.65];0.0001*** | 0.56[0.42-0.74];<0.0001*** | 0.67[0.60-0.76];<0.0001*** |
| ***Model 6*** | 0.45[0.30-0.70];0.0003*** | 0.57[0.40-0.76];<0.0001*** | 0.68[0.46-0.78];<0.0001*** |

**Supplementary Table 3. Sensitivity analyses for exposure effects of SGLT2I v.s. DPP4I on new onset prostate cancer, cancer-related mortality, and all-cause mortality using different models.**
* for p≤ 0.05, ** for p ≤ 0.01, *** for p ≤ 0.001; SGLT2I: Sodium-glucose cotransporter-2 inhibitors; DPP4I: Dipeptidyl peptidase-4 inhibitors; HR: hazard ratio; CI: confidence interval; PS: propensity score; IPTW: inverse probability of treatment weighting, SIPTW: stable inverse probability of treatment weighting.

| **Model** | **New onset prostate cancer HR [95% CI];P value** | **Cancer-related mortality HR [95% CI];P value** | **All-cause mortality HR [95% CI];P value** |
|---|---|---|---|
| Cause-specific hazard models | 0.31[0.23, 0.41];<0.0001*** | 0.24[0.19, 0.30];<0.0001*** | 0.21[0.19, 0.23];<0.0001*** |
| Sub-distribution hazard models | 0.45[0.33, 0.56];<0.0001*** | 0.35[0.22, 0.41];<0.0001*** | 0.34[0.20, 0.44];<0.0001*** |
| PS stratification | 0.45[0.26, 0.56];<0.0001*** | 0.36[0.26, 0.51];<0.0001*** | 0.34[0.22, 0.49];<0.0001*** |
| PS with IPTW | 0.56[0.34, 0.62];<0.0001*** | 0.45[0.34, 0.58];<0.0001*** | 0.41[0.28, 0.55];<0.0001*** |
| PS with SIPTW | 0.58[0.33, 0.59];<0.0001*** | 0.46[0.33, 0.61];<0.0001*** | 0.46[0.23, 0.59];<0.0001*** |

**Supplementary Table 4. Sensitivity analysis: Three-arm (only SGLT2I, only DPP4I, and only GLP1a) analysis results using stabilized IPTW.**

* for p≤ 0.05, ** for p ≤ 0.01, *** for p ≤ 0.001; HR: hazard ratio; CI: confidence interval; SGLT2I: sodium glucose cotransporter-2 inhibitor; DPP4I: dipeptidyl peptidase-4 inhibitor; glucagon-like peptide-1 receptor agonist (GLP1a)

| **New onset prostate cancer** | **HR [95% CI]** | **P value** |
|---|---|---|
| DPP4I v.s. SGLT2I | 1.80 [1.26-2.58] | 0.0013*** |
| GLP1a v.s. SGLT2I | 1.15 [0.84-1.58] | 0.3931 |
| **Cancer related mortality** | **HR [95% CI]** | **P value** |
| DPP4I v.s. SGLT2I | 2.44 [1.90-3.12] | <0.0001*** |
| GLP1a v.s. SGLT2I | 1.07 [0.48-2.41] | 0.8642 |
| **All-cause mortality** | **HR [95% CI]** | **P value** |
| DPP4I v.s. SGLT2I | 2.63 [2.36-2.92] | <0.0001*** |
| GLP1a v.s. SGLT2I | 1.01 [0.68-1.50] | 0.9633 |

**Supplementary Table 5. Sensitivity analysis: Excluding patients with CKD stage 4/5 (eGFR <30), peritoneal dialysis or haemodialysis in the SGLT2I v.s. DPP4I matched cohort.**

* for p≤ 0.05, ** for p ≤ 0.01, *** for p ≤ 0.001; SGLT2I: Sodium-glucose cotransporter-2 inhibitors; DPP4I: Dipeptidyl peptidase-4 inhibitors; HR: hazard ratio; CI: confidence interval

| | **All-cause mortality HR [95% CI];P value** | **Cancer-related mortality HR [95% CI];P value** | **New onset prostate cancer HR [95% CI];P value** |
|---|---|---|---|
| SGLT2I v.s. DPP4I | 0.60[0.53-0.68];<0.0001*** | 0.51[0.38-0.68];<0.0001*** | 0.47[0.30-0.76];0.0018** |

**Supplementary Table 6. Sensitivity analysis: Consideration of 1-year lag time effects in the SGLT2I v.s. DPP4I matched cohort.**

* for p≤ 0.05, ** for p ≤ 0.01, *** for p ≤ 0.001; SGLT2I: Sodium-glucose cotransporter-2 inhibitors; DPP4I: Dipeptidyl peptidase-4 inhibitors; HR: hazard ratio; CI: confidence interval

| | **All-cause mortality HR [95% CI];P value** | **Cancer-related mortality HR [95% CI];P value** | **New onset prostate cancer HR [95% CI];P value** |
|---|---|---|---|
| SGLT2I v.s. DPP4I | 0.51[0.46-0.57];<0.0001*** | 0.36[0.28-0.45];<0.0001*** | 0.45[0.29-0.69];0.0003*** |
